# DNA methylation signatures and the contribution of age-associated methylomic drift to carcinogenesis in early-onset colorectal cancer

**DOI:** 10.1101/2021.03.24.21254210

**Authors:** Jihoon E. Joo, Mark Clendenning, Ee Ming Wong, Christophe Rosty, Khalid Mahmood, Peter Georgeson, Ingrid M. Winship, Susan G. Preston, Aung Ko Win, Pierre-Antoine Dugué, Harindra Jayasekara, Dallas English, Finlay A. Macrae, John L. Hopper, Mark A. Jenkins, Roger L. Milne, Graham G. Giles, Melissa C. Southey, Daniel D. Buchanan

## Abstract

**Background:** The role of DNA methylation (DNAm) in the carcinogenesis of colorectal cancer (CRC) diagnosed <50years of age (early-onset CRC or EOCRC) is currently unknown. We investigated aberrant DNAm changes and the contribution of ageing-associated methylomic drift, and age acceleration to EOCRC carcinogenesis.

**Methods:** Genome-wide DNAm profiling using the Infinium HM450K on 97 EOCRC tumour and 54 normal colonic mucosa samples was compared with: 1) intermediate-onset CRC (IOCRC; diagnosed between 50-70 years; 343 tumour and 35 normal); and 2) late-onset CRC (LOCRC; >70 years; 318 tumour and 40 normal). CpGs associated with age-related methylation drift were identified using a public dataset of 231 normal mucosa samples from people without CRC. DNAm-age was estimated using epiTOC2.

**Results:** Common to all three age-of-onset groups, 88,385 (20% of all CpGs) CpGs were differentially methylated between tumour and normal mucosa. We identified 234 differentially methylated genes that were unique to the EOCRC group. In normal mucosa from people without CRC, we identified 28,154 CpGs that undergo ageing-related DNAm drift and of those, 65% were aberrantly methylated in EOCRC tumours. Based on the mitotic-based DNAm clock epiTOC2, we identified age acceleration in normal mucosa of people with EOCRC compared with normal mucosa from the IOCRC, LOCRC groups (*p*=3.7×10^−16^) and young people without CRC (*p*=5.8×10^−6^).

**Conclusion:** EOCRC acquires unique DNAm alterations at 234 loci. CpGs associated with ageing-associated drift were widely affected in EOCRC without needing the decades-long accrual of DNAm drift as commonly seen in intermediate- and late-onset CRCs. We found accelerated ageing in normal mucosa from people with EOCRC, as evidenced by a faster stem-cell division rate, potentially contributing to EOCRC carcinogenesis.

## Introduction

Colorectal cancer (CRC) is the third most commonly diagnosed cancer and the second leading cause of cancer-related death worldwide (1). CRC is a heterogeneous disease, for which increasing age is one of the strongest risk factors (1). Screening programs have been implemented for people starting at age 50 years in many countries, which has contributed to a reduction in CRC incidence in this older age group (2). In contrast, the incidence rate of CRC in people under 50 years of age has been increasing and now accounts for over 10% of all CRC diagnoses in the US (3). Although monogenic cancer predisposition syndromes may explain up to 20% of all early-onset colorectal cancer (EOCRC) (4,5), it is unlikely to explain the increasing incidence of EOCRC. Currently, we do not know if EOCRC is molecularly distinct from later-onset CRC, a knowledge gap that hinders efforts to identify the cause/s of this increasing incidence.

CRCs acquire aberrant DNA methylation (DNAm), including focal hypermethylation leading to silencing of tumour suppressor genes and global hypomethylation resulting in genomic instability (6). Global hypomethylation in tumour and hypermethylation in blood have been shown to be more frequent in EOCRC when compared with non-hereditary CRCs diagnosed at age >50 years (7,8). To date, there has been no study exploring the aetiologic role of genome-wide DNA methylation changes in EOCRC. DNAm can be modified by environmental exposures, such as diet and is likely to be a primary mechanism explaining environmentally-modulated EOCRC risk (9). Further, DNAm changes at both cancer-related and ageing-related CpG loci have been identified in normal colonic mucosa (NM), suggesting that DNAm alterations may start very early in CRC tumorigenesis and predispose cells to neoplastic transformations (10-12), hence they are a promising biomarker for predicting CRC risk (6).

Tissue-specific DNAm changes, including both hyper- and hypomethylation of CpGs, increase as part of the normal ageing process, a phenomenon referred to as “methylomic drift” (13). The decades-long accumulation of methylomic drift is thought to underlie the association between ageing and CRC risk (13,14), where adverse endogenous and environmental stimuli can accelerate the rate of this methylomic drift (13,15). Furthermore, DNAm is shown to be an accurate molecular indicator for biological age and this has led to the development of numerous tools for estimating biological age using genome-wide DNAm data (16-22). These tools have been used to investigate the difference between chronological and biological age, referred to as age acceleration (AA), and AA is associated with several diseases, including cancer risk (18).

Current evidence suggests that methylomic drift occurs more rapidly in colorectal neoplasia than in NM but that cancer precursors frequently sojourn for decades before transitioning into cancer, implying that the founder premalignant cell typically arises early in life (14). We hypothesise that EOCRC is characterised by specific DNAm aberrations including those related to rapid methylomic drift and AA compared with later-onset CRC. In this study, we assessed genome-wide DNAm in tumour and NM samples from people with EOCRC (CRC diagnosed at age/AgeDx ≤50 years) as well as in people with intermediate-onset CRC (IOCRC; AgeDx between 50-70 years) and with late-onset CRC (LOCRC; AgeDx > 70 years). Our aims were to determine: i) the molecular uniqueness of EOCRC tumours related to DNAm aberrations and the affected gene pathways and ii) the role of methylomic drift and AA in the NM and tumours from people with EOCRC compared with people with LOCRC.

## Materials and Methods

### Study participants

People affected with CRC were identified through the Australian Colorectal Cancer Family Registry (ACCFR) (23,24) and Melbourne Collaborative Cohort Study (MCCS) (25). The ACCFR recruited people diagnosed with CRC at <60 years of age identified by linkage to the Victorian Cancer Registry between 1998 and 2008. CRC-affected participants diagnosed at <50 years of age with available CRC tissue and matched normal colonic mucosa were selected for testing in this study. The MCCS is a prospective cohort study with 41,513 participants from the Melbourne metropolitan area with a mean age of 55 years when recruited between 1990 and 1994 (25). By 31 December 2009, 1,046 participants had a first histopathological diagnosis of invasive adenocarcinoma of the colon or rectum identified by record linkage to the Victorian Cancer Registry following the baseline study visit. Germline and tumour characterisation of these CRC-affected probands from both studies has been previously described (26). In total, CRC tumour and matched NM samples (where available) from 769 individuals were included in the analysis. Individuals from both studies who carried a germline mutation in one or more of the DNA mismatch repair (MMR) genes (i.e. *MLH1, MSH2, MSH6* and *PMS2*) or the *MUTYH* gene were excluded from the analysis. In addition, only CRCs that showed normal and retained expression of all four MMR proteins by immunohistochemistry (i.e. MMR-proficient) were included in the analysis. For all subsequent analyses, samples were divided into three groups based on age at diagnosis (AgeDx): 1) ***Early* or EOCRC** (CRC AgeDx ≤ 50 years), 2) ***intermediate* or IOCRC** (AgeDx between 51 and 70 years) and 3) ***late-*onset or LOCRC** (AgeDx >70 years).

### DNA extraction from FFPE specimens

Formalin-fixed-paraffin-embedded (FFPE) tissue specimens from the tumour and matched NMs from the surgical resection margin were identified. All haematoxylin & eosin slides were reviewed by specialist GI pathologists and regions enriched for normal or tumour cells were marked up for macrodissection. DNA was extracted using QIAamp DNA FFPE tissue kit (Qiagen, Germany) and quantified using the Qubit dsDNS HS kit (Thermo Scientific, Carlsbad, CA). Up to 500ng of genomic DNA isolated from FFPE slides was bisulfite converted using the EZ DNA Methylation-Gold Kit (Zymo Research, CA), following the manufacturer’s instruction. Bisulfite converted DNA was restored using the FFPE restore kit (Illumina, CA).

### DNA methylation array processing and bioinformatic analyses

Genome-wide DNAm was profiled using the Infinium HumanMethylation450K array (HM450K; Illumina) and processed as previously described (27). The staining and extension steps were performed using the TECAN EVO automated liquid handler (Männedorf, Switzerland). Raw data were imported into R programming software (v3.3.2) and processed using the *minfi* Bioconductor package (28). Raw intensity data were converted into methylset and normalised using *Functional normalisation* (29) with *noob* background correction (30), which are implemented in *minfi* (28). Samples with mean detection p-values greater than 0.01 were removed from the analysis. Probes with detection p-value greater than 0.05 were deemed noisy and excluded from the analysis. β (Beta) and M-values were calculated using *getBeta* and *getM* function in *minfi*. β-values were used for presenting the data and M-values were used for all statistical analyses as suggested previously (31). To eliminate any sex-specific bias in DNA methylation, probes on sex chromosomes were removed from all analyses and methylation levels were measured from 431,942 probes in total.

### Statistical analysis

Differentially methylated probes (DMPs) were identified using the *limma* Bioconductor package (32). Differentially methylated regions (DMRs) were identified using the *DMRcate* Bioconductor package (33). All p-values were FDR-adjusted for multiple testing unless stated otherwise. The differential methylation analyses between tumour and matched NMs (from same individuals) were performed in a pairwise design. All plots were generated using *ggplot*2 R packages (34). All statistical analyses were performed in R. KEGG pathway analyses were performed on DMRs using the *gometh* function from the *MissMethyl* Bioconductor package (35).

### Publicly available DNA methylation dataset of normal colonic mucosa and CRCs

CpGs associated with methylomic drift were identified using a publicly available methylation dataset of 231 NM samples collected from healthy participants without CRC from SMS and CICaRes cohorts (GEO Accession “GSE113904”) (14,36,37), which was downloaded using the *minfi* package (28). One sample was removed due to missing anatomical site information. Raw “Drift-CpGs” were identified by performing a regression analysis between M-values of individual CpG sites assessed by the HM450K and the age in years, which was fit using splines.

### DNA methylation-based age estimation

To calculate DNAm-based biological age (DNAmAge), we pooled the healthy NM data (n =231) (14), with our NMs (n = 129) and CRC (n = 758) data. The pooled data underwent a *ComBat* batch correction (38) by treating individual slides (or “chips”) as a batch (**Supplementary Figure 1**). DNAmAge was assessed using five well-established algorithms. The Horvath clock was calculated via https://genetics.ucla.edu/new (17). The Hannum age (16) and PhenoAge (19) were estimated using the *ENmix* Bioconductor package (39). The mitotic age was assessed by epiTOC (or “pcgtAge”) (21) and epiTOC2 (or “tnsc2”), which were calculated using the method and the R code detailed in Teschendorff et al (20). The irS (average intrinsic rate of stem-cell division per year) was also derived from the R code by inputting corresponding samples’ chronological ages. Age acceleration (AA) was determined by estimating residuals from a linear regression of DNAmAge on chronological age, and by adjusting for sex and cell heterogeneity (where relevant) as previously demonstrated (17,18,40,41). For the Horvath clock, residual AA (“AgeAccelerationResidual”) was derived from the online calculator by uploading chronological age, gender and tissue source (17).

### Ethics approval

This study was approved by Cancer Council of Victoria and University of Melbourne Human Research Ethics Committees.

## Results

In total, we analysed genome-wide DNAm data from 758 CRC tumour and 129 histologically normal mucosa (NM) samples from 769 individuals stratified into EOCRC, IOCRC and LOCRC subgroups (**Table 1**). Characteristics of the participants and their tumours are described in **Table 1**. A principal component analysis of all detected probes showed that samples clustered by tissue type and no batch effect was observed (**Supplementary Figure 2**).

**Table 1.** CRC-affected participant and tumour characteristics for early, intermediate and late-onset groups.

|  | *Onset of disease* | **Early (EOCRC)** |  | **Intermediate (IOCRC)** |  | **Late (LOCRC)** |  |
| --- | --- | --- | --- | --- | --- | --- | --- |
|  | *Ages of onset* | < 50 years |  | ≥ 50 & <70 years |  | ≥ 70 years |  |
|  | *Mean (sd)* | 36.2 (6.2) years |  | 62.6 (5.2) years |  | 74.8 (3.5) years |  |
|  | *Total number of cases*<br>*(matched tumour/normal pairs)* | 110 (41) |  | 334 (34) |  | 325 (27) |  |
|  | *Sex (%)* | F | M | F | M | F | M |
|  |  | 64 (58) | 46 (42) | 158 (47) | 176 (53) | 142 (43) | 183 (57) |
|  | *Tissue type* | Tumour | Normal | Tumour | Normal | Tumour | Normal |
|  |  | 97 | 54 | 343 | 35 | 318 | 40 |
| *Anatomical site (%)* | *Right colon* | 20 (20.5) | 11 (21) | 86 (25) | 8 (23) | 107 (34) | 14 (35) |
|  | *Left colon* | 32 (33.5) | 13 (24) | 100 (29) | 8 (23) | 91 (29) | 9 (22.5) |
|  | *Rectum* | 40 (41) | 19 (35) | 138 (40) | 8 (23) | 114 (36) | 7 (17.5) |
|  | *unknown* | 5 (5) | 11 (20) | 19 (6) | 11 (31) | 6 (2) | 10 (25) |
| *TNM (AJCC) stage (%)* | *Stage I* | 17 (18) | - | 70 (20) | - | 75 (24) | - |
|  | *Stage II* | 25 (26) | - | 81 (24) | - | 81 (25) | - |
|  | *Stage III* | 43 (44) | - | 101 (30) | - | 87 (27) | - |
|  | *Stage IV* | 9 (9) | - | 54 (16) | - | 55 (17) | - |
|  | *unknown* | 3 | - | 37 | - | 20 | - |
| *CIMP group* | *CIMP +* | 0 | - | 19 | - | 34 | - |
|  | *CIMP -* | 93 | - | 320 | - | 280 | - |
|  | *unknown* | 4 | - | 4 | - | 4 | - |

### Differences in genome-wide DNA methylation between normal mucosa and CRCs by age-of-diagnosis

The pairwise analysis of matched CRC and NM samples within the three defined age groups identified differentially methylated probes (DMPs): 29% of all (431,942) CpGs in 41 EOCRCs, 34% in 34 IOCRCs and 30% DMPs in 27 LOCRCs (**Figure 1** & **Table 2**). These DMPs were associated with 17,666, 20,221 and 17,769 differentially methylated ***regions*** or ***genes*** (DMRs) in EOCRC, IOCRC and LOCRC groups, respectively. Hypomethylated DMRs represented 60%, 61% and 53% of the overall DMRs in the EOCRC, IOCRC and LOCRC groups, respectively (**Table 2**) and the difference in proportions was statistically significant (*p <* 0.001).

**Table 2.** Numbers of differentially methylated probes and regions identified between tumour and normal pairs from EOCRC, IOCRC and LOCRC cases included in the study. ^#^ Pearson’s Chi-squared test

|  |  | *Early-onset*<br>*(EOCRC)* | *Intermediate-onset*<br>*(IOCRC)* | *Late-onset*<br>*(LOCRC)* | *P-value* |
| --- | --- | --- | --- | --- | --- |
| *Age of Dx* |  | *< 50 years* | *≥ 50 & <70 years* | *≥ 70 years* |  |
| *N of tumour and matched normal pairs* |  | *41* | *34* | *27* |  |
| *Number of DMPs in total*<br>*(FDR adj. P < 0.01)* | *Hypermethylated* | *49,044 (38.8%)* | *59,125 (40.3%)* | *57,970 (45.1%)* | *< 0.001<sup>#</sup>* |
|  | *Hypomethylated* | *77,228 (61.2%)* | *87,448 (59.7%)* | *70,806 (54.9%)* |  |
|  | *Total* | *126,272 (29.2% of all CpGs included in the analysis)* | *146,573 (33.9%)* | *128,776 (29.8%)* |  |
| *Number of DMRs\* (FDR adj. P < 0.01)* | *Hypermethylated* | *7,102 (40%)* | *7,985 (39.5%)* | *8,407 (47.3%)* | *< 0.001<sup>#</sup>* |
|  | *Hypomethylated* | *10,564 (60%)* | *12,236 (60.5%)* | *9,362 (52.7%)* |  |
|  | *Total* | *17,666* | *20,221* | *17,769* |  |
| *Number of DMPs that are only presented in each group (FDR adj. P < 0.01)* | *Hypermethylated* | **7,369 (39.3%)** | **9,171 (40%)** | **10,216 (69%)** | **< 0.001<sup>#</sup>** |
|  | *Hypomethylated* | **11,386 (60.7%)** | **13,727 (60%)** | **4,581 (31%)** |  |
|  | *Total* | **18,755 (4.3%)** | **22,898 (5.3%)** | **14797 (3.4%)** |  |
| *Number of DMRs that are only presented in each group\* (FDR adj. P < 0.01)* | *Hypermethylated* | **932 (42.2%)** | **1,459 (53%)** | **1,336 (84%)** | **< 0.001<sup>#</sup>** |
|  | *Hypomethylated* | **1,276 (57.8%)** | **1,298 (47%)** | **254 (16%)** |  |
|  | *Total* | **2,208** | **2,757** | **1,590** |  |

**Figure 1.**
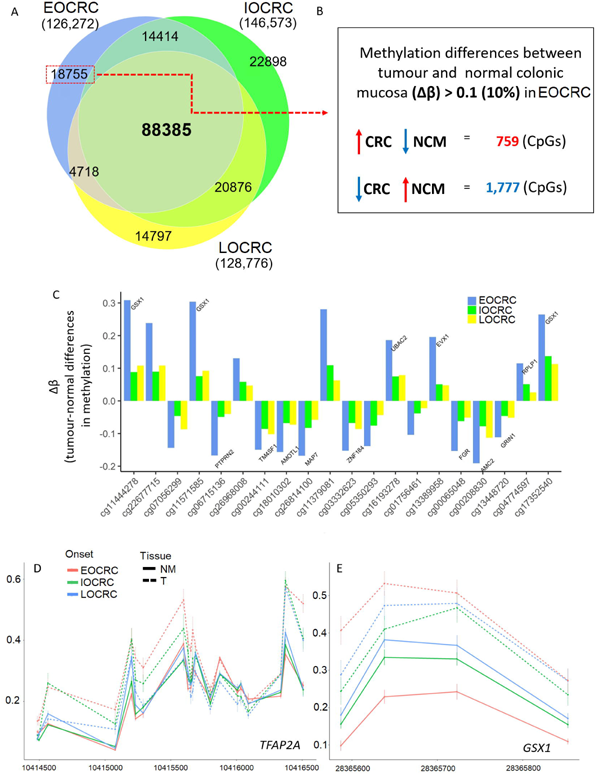
Differential methylation between matched tumour and normal pairs for three age groups. **A)** (Proportional) Venn diagram showing the numbers of differentially methylated probes (DMPs) between tumour and matched normal mucosa samples for 41 EOCRCs, 34 IOCRCs and 27 LOCRCs. **B)** Of 18,755 DMPs with a Δβ greater than 0.1 that were unique to the tumour-normal pairs from EOCRC, 759 DMPs were hypermethylated and 1,777 DMPs were hypomethylated. **C)** Barplot illustrating differences in methylation levels (Δβ) between tumour and normal pairs from EOCRC (blue), IOCRC (green) and LOCRC (yellow) for the top 20 DMPs ranked by statistical significance (genes that overlap DMPs are shown). **D&E**. DNA methylation patterns two highly ranked DMRs (**D** – *TFAP2A*, chr6:10414350– 10416531 & **E –** *GSX1*, chr13:28365587-28365853) where greater methylation differences are specific to samples from EOCRC.

We identified 88,385 DMPs that overlapped between the three CRC-onset groups, however, each age group also showed a subset of unique DMPs (**Table 2 & Figure 1A**). Of 18,755 DMPs that were unique to the EOCRC group, 39% were hypermethylated in CRCs when compared with matched NM samples. This proportion of hypermethylated CpGs was different to IOCRC and LOCRC (**Table 2**). When restricting to DMPs showing large changes in methylation (absolute mean Δβ values > 0.1), the number of EOCRC DMPs was reduced to 2,536 DMPs with 759 (30%) hypermethylated and 1,777 (70%) hypomethylated CpGs in tumours (**Figure 1B** & **Figure 1C**). A kernel smoothing of these 2,536 CpGs revealed 234 DMRs (**Supplementary Table 1**). The top-ranked DMRs (by Stouffer’s *p-* value (33)) included: 1) the transcription start site (∼2kb) of the *TFAP2A* gene (**Figure 1D**), 2) a short intergenic region on chromosome 1, and 3) a ∼200bp region located ∼1kb upstream of the *GSX1* gene (**Figure 1E)**.

Restricting to DMRs with absolute mean differences in methylation (Δβ) > 0.1 to remove genes with subtle methylation changes which are less likely to impact gene function, we performed the KEGG analysis to examine biological pathways associated with DMRs for each age group. For the EOCRC group, 5,585 DMRs (**Supplementary Table 2**) were associated with 10 KEGG pathways (FDR-adjusted *p* < 0.05) (**Supplementary Figure 3** & **Supplementary Table 3**). For the IOCRC and LOCRC groups, 12 and 18 KEGG pathways (**Supplementary Table 4** & **Supplementary Table 5**) were associated with 7,701 and 7,928 DMRs (**Supplementary Table 6** & **Supplementary Table 7**), respectively. Eight KEGG pathways were common to all three age groups including “Neuroactive ligand-receptor interaction” pathway which was the most significant pathway for all three onset groups. The “Maturity onset diabetes of the young” pathway was uniquely associated with the DMRs from the EOCRC group.

### Methylomic drift in normal colonic mucosa and colorectal carcinogenesis

We identified CpGs at which DNAm levels consistently change or “drift” with ageing using a previously published dataset of 231 NM samples collected from only people described as “healthy”, unaffected by CRC, ranging in age from 29 to 81 years [mean = 59 ± 10 s.d.] (14,36,37). After adjusting for anatomical site (proximal and distal colon and rectum), the analysis revealed 30,465 drift-CpGs (*FDR adj P <* 0.01) (**Figure 2 & Supplementary Table 8**). The majority (29,085 CpGs or 96%) were positively correlated (Spearman’s ranked correlated coefficient _ρ_ > 0) with age (**Figure 2**). In the 129 NM samples from people with CRC, the association between methylation levels and age was weaker (**Figure 2B**) when tested on 28,154 drift-CpGs (excluding 2,311 poorly performing probes from our data).

**Figure 2.**
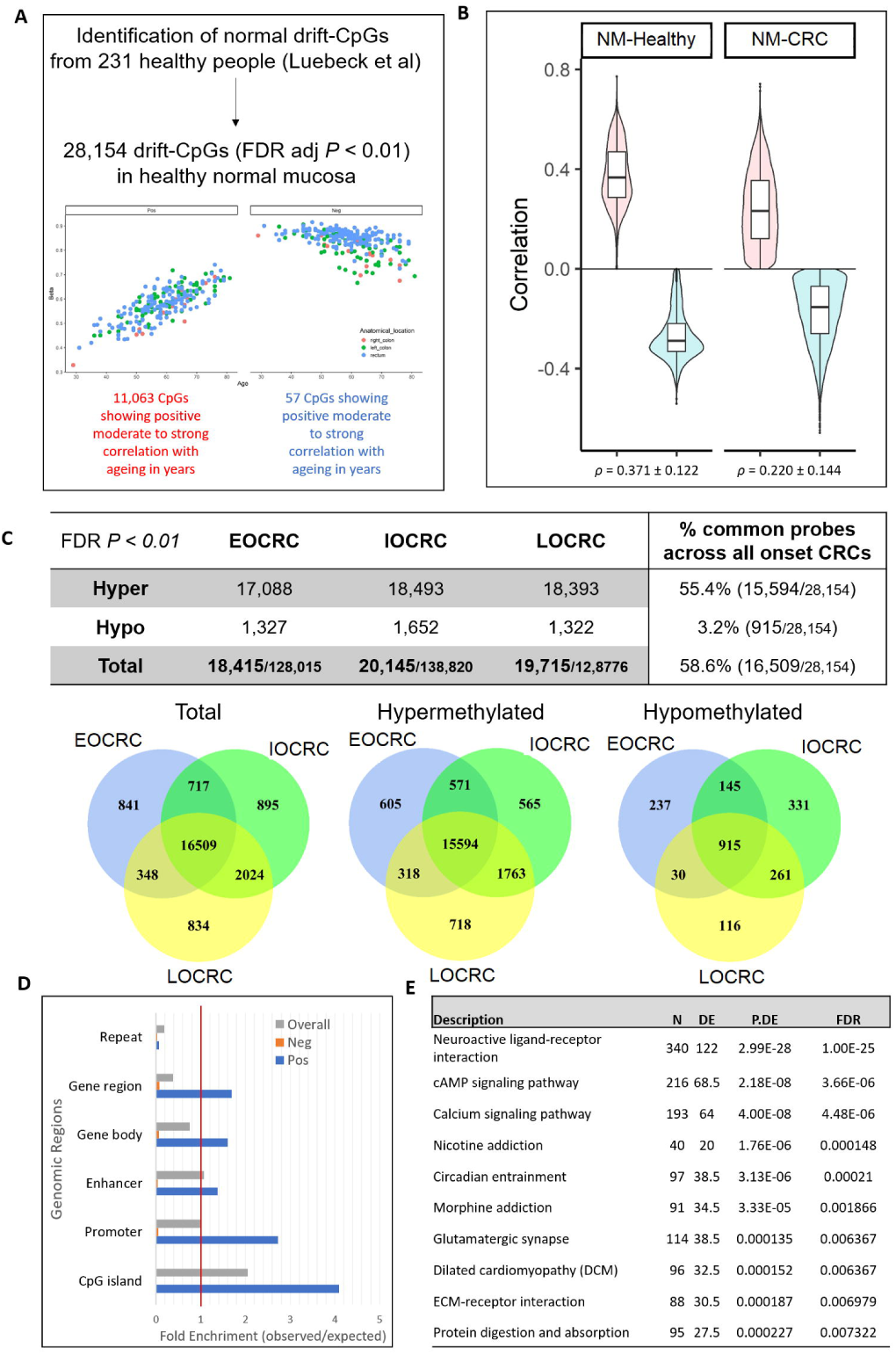
CpGs associated with the methylomic drift in normal colonic mucosa and those that are also DMPs in CRCs **(A)** 28,154 CpGs were identified where methylation levels were either positively or negatively correlated with increasing age i.e. “drifts-CpGs”. **(B)** Spearman’s correlation coefficients for 26,886 positively correlated probes (red) and 1,268 negatively correlated probes (blue) in normal mucosa from healthy control (“NM-healthy”, left) and people with CRC from this study (“NM-CRC”, right). (**C**) A table and venn diagrams showing the breakdown of hypermethylated and hypomethylated cancer-associated drift-CpGs each for EOCRC, IOCRC and LOCRC. (**D**) The representation of genomic regions (CpG island, gene promoter, enhancer, gene body and repeat) for 16,509 cancer-associated drift CpGs common between all three age groups that were either positively or negatively correlated drift-CpGs. (**E**) Top 10 KEGG pathways associated with 16,509 cancer associated drift CpGs.

We next measured the overlap between the 28,154 drift-CpGs and the DMPs identified from our three CRC age groups. The majority of these drift-CpGs were aberrantly methylated in tumours, with 65%, 72% and 70% of the 28,154 drift-CpGs overlapping with DMPs from the EOCRC, IOCRC and LOCRC groups, respectively (**Figure 2C**). Intriguingly, these cancer-associated drift CpGs (“CA-drift CpGs”) were broadly affected even in EOCRC, with 59% (16,509/28,154) common across all three age groups. The CA-drift CpGs clustered into 2,824 DMRs linked to 2,597 annotated genes and 227 intergenic regions (**Supplementary Table 9**).

Almost all 15,982 CA-drift CpGs, for which methylation was positively correlated with ageing were hypermethylated (15,578 CpGs or 97%) in tumours and this pattern was consistent across the three age-of-diagnosis groups. The CA-drift CpGs were located largely within intragenic and regulatory regions including gene promoter and enhancer regions, while CpGs within repeat and gene body regions were underrepresented (**Figure 2D**). A KEGG pathway analysis of the 16,509 CA-drift CpGs identified several biological pathways associated with genes overlapping those CpG sites, as shown in **Figure 2E**.

Of the 28,154 drift-CpGs identified, 3% (841/28,154 CpGs) overlapped with cancer DMPs that were unique to EOCRC (**Figure 3A**), which clustered into 86 regions (80 annotated genes and 6 intergenic) (**Supplementary Table 10**). The *GSX1* and *TFAP2A* regions, mentioned in the above analysis, were among the highly ranked CA-drift CpGs specific to EOCRC. Other genes associated with CA-drift CpGs of EOCRC included *PRRT1, PHF11* and *ZFP42* (**Supplementary Table 10**). We observed enrichment of “Systemic lupus erythematosus” (*FDR-*adj. *p* = 0.0002) and “Alcoholism” (*FDR*-adj *p =* 0.004) KEGG pathways associated with the 86 regions.

**Figure 3.**
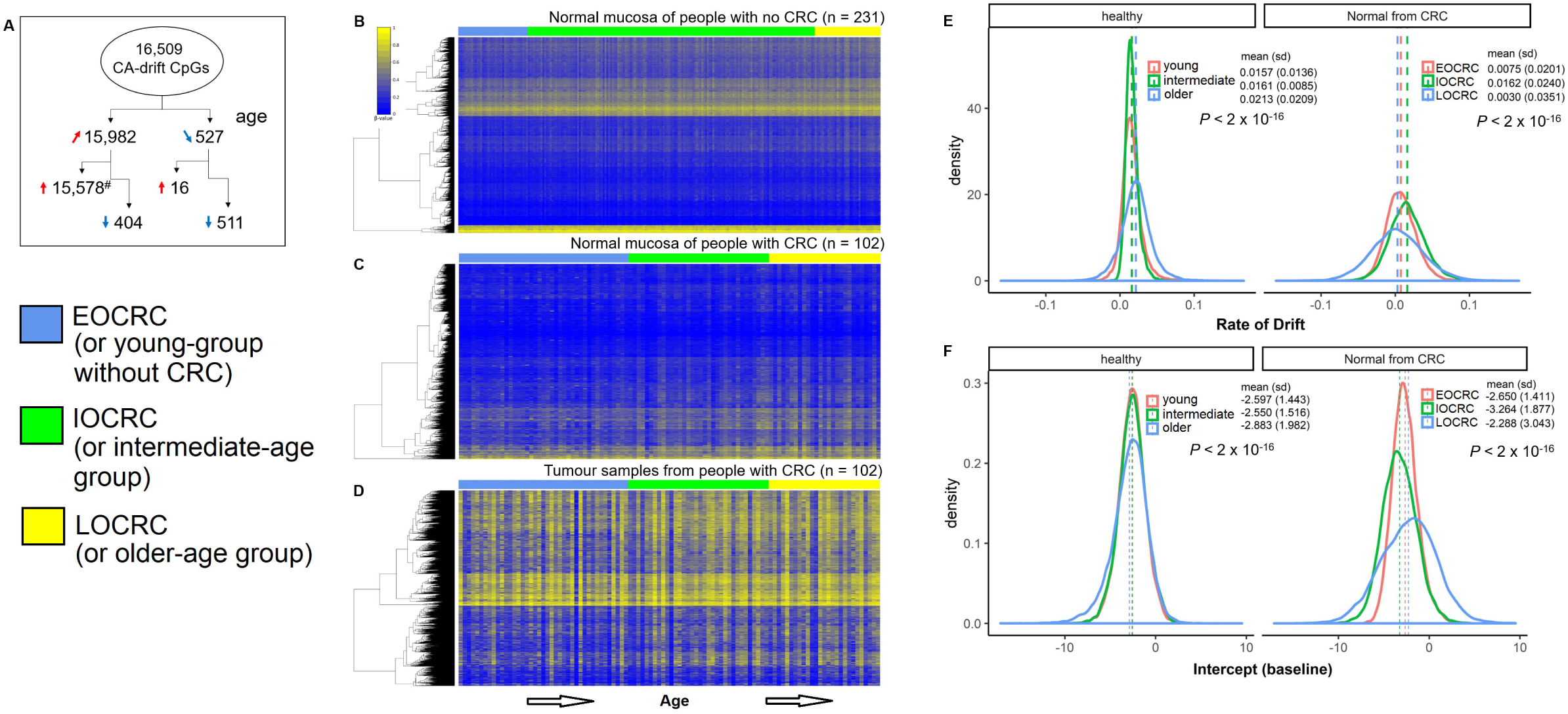
**(A)** Schema describing number breakdowns for CA-drift CpGs positively and negatively correlated with increasing age and hyper and hypomethylated in CRCs. (**B)** DNA methylation heatmap illustrating methylation patterns across 15,578 CA-drift CpGs (y-axis) for 231 normal mucosa samples (x-axis) from the healthy group. Individual samples are shown along the horizontal axis and order by increasing age. **(C)** for 102 normal mucosa samples from people with CRC. **(D)** for 102 tumour samples from people with CRC. **(E)** Density plots showing the distribution of the drift rate for young/early, intermediate and late age group samples from people without CRC and with CRC. **(F)** Density plots showing the baseline of the drift rate (using intercept as surrogate).

### Rate of change of DNA methylation at cancer-associated drift CpGs

DNAm changes (or drift) at the CA-drift CpGs occurred slowly in NMs from both the people without CRC (mean changes in β/10yrs = 0.02 ± 0.010 s.d.) and people with CRC (mean changes in β/10yrs = 0.007 ± 0.014 s.d.) (**Figure 3B&C**) and this difference was statistically significant (*p* < 0.001). In contrast, a substantial degree of DNAm changes at CA-drift CpGs were observed in CRC tumour samples irrespective of age of diagnosis (**Figure 3D**). We evaluated the drift rate at individual 15,578 CpGs by fitting a linear model between the age and M-values for individual CA-drift CpGs as previously described (14).

Small but statistically significant differences in the pattern of the drift rates between the three age-of-diagnosis groups were observed for NMs from both healthy and CRC affected groups (*p* <0.001, **Figure 3E**). Whilst the differences in mean drift rates did not show strong differences between the three age groups, the variations were notably large (*p* <0.001) in the later age groups in NMs from both healthy and CRC-affected groups (**Figure 3E**). This trend between the three age groups were also significantly different between NMs from people with or without CRC (*P* < 0.001), due to the larger variations in the drift rate for the CRC group overall (**Figure 3E**). We also estimated the baseline of the drift for individual groups using the intercept from the linear model as a surrogate. Similarly, we observed small but significant differences between the three age groups but not between groups with CRC and without CRC (**Figure 3F**).

### DNA methylation-based biological age of the normal mucosa and age acceleration

We assessed the “epigenetic age” of each tissue sample using five well-established DNAm-based clock algorithms (16,17,19), including two recently developed “mitotic-like” clocks, epiTOC (21) and epiTOC2 (20). These two mitotic-like DNAm clocks have demonstrated their effectiveness in discriminating precancerous from normal tissues, showing utility for predicting cancer risks from pre-diagnostic tissue samples (20,21). Unlike non-mitotic DNAm clocks, these clocks quantify the magnitude of DNAm errors that arose throughout every stem cell division by interrogating DNAm levels across the promoters of polycomb-associated genes (20,21).

By both epiTOC and epiTOC2, moderate correlations (r = 0.462 and 0.476, respectively) were observed between DNAm age and chronological age in NMs from people without CRC, whereas this was notably poorer in NMs from people with CRC (r = 0.030 and 0.054, respectively) (**Figure 4A & B**). In CRCs, weak correlations (r = 0.284 and 0.274 for epiTOC and epiTOC2, respectively) were observed but with greater, and a broader range of, DNAm ages compared with the two NM groups. All three non-mitotic clocks (i.e. Horvath (17), Hannum (16) and PhenoAge (19)) generally showed moderate-to-strong correlations in NMs, as previously demonstrated (41) but not in CRCs (**Supplementary Figure 4**).

**Figure 4.**
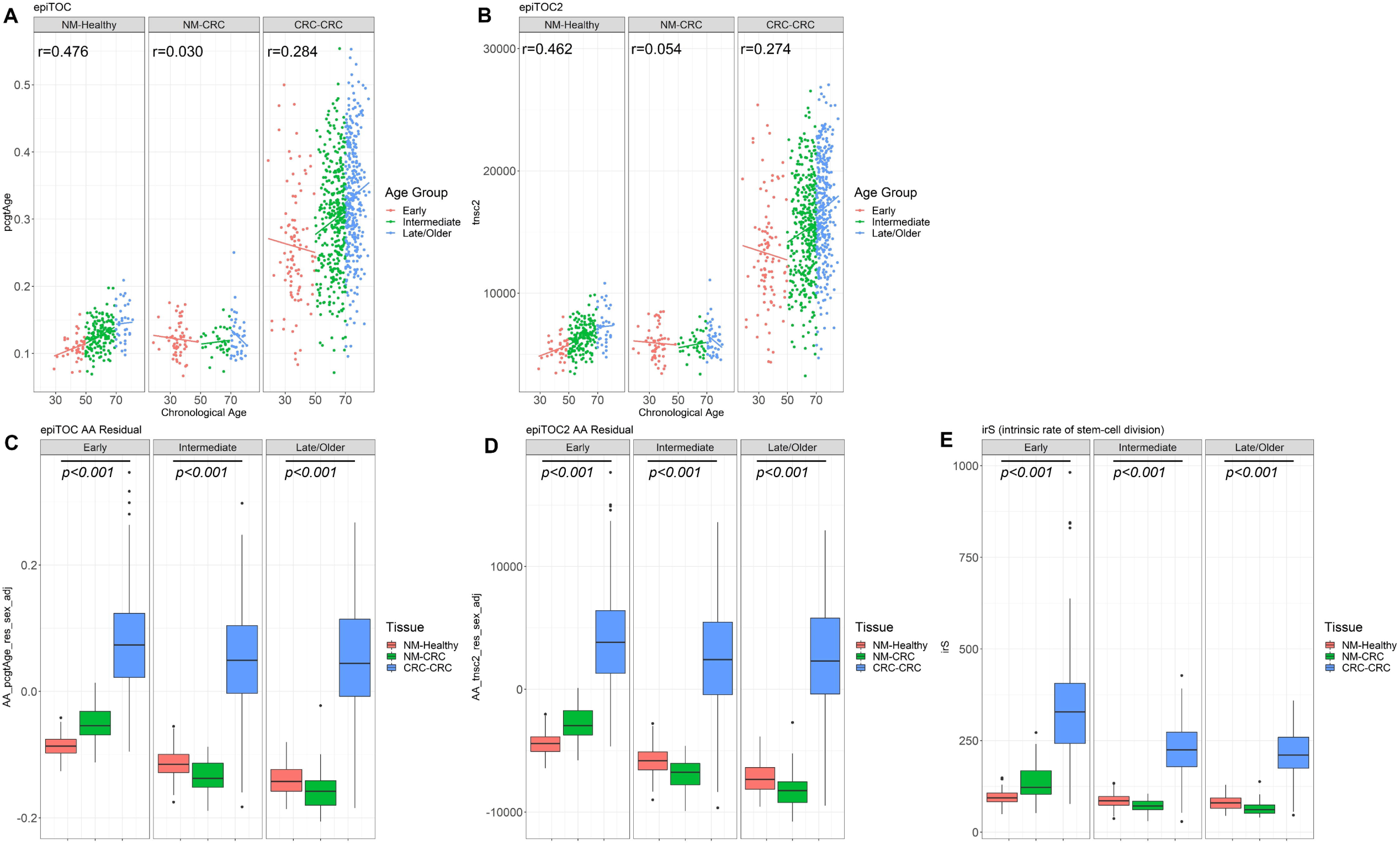
The mitotic-based DNAms age prediction by epiTOC and epiTOC2 for normal colonic mucosa samples from people without (“**NM-Healthy”**) and people with CRC (“**NM-CRC”**), and CRC tumour (“**CRC-CRC”**) samples, separated into **Early, Intermediate** and **Late/Older** age groups based on the age at tissue collection or CRC diagnosis. **A**. Scatterplots illustrating correlations between chronological age (x-axis) and epiTOC DNAm-based age (y-axis) for corresponding samples. **B**. Scatterplots illustrating correlation between chronological age and epiTOC2 DNAm-based age (y-axis). Boxplots illustrating the distribution of age-acceleration (AA) as estimated using epiTOC (**C**), epiTOC2 (**D**) and irS (average lifetime intrinsic rate of stem-cell division per sample), as derived from epiTOC2 (**E**), shown separately by the age and the tissue groups. *P*-values were obtained from Kruskal-Wallis Rank Sum tests.

Using the five epigenetic clocks, we assessed AA by estimating residuals by regressing DNAm (or mitotic) ages onto the chronological ages for all samples (40,41), adjusting for sex. Additionally, the average intrinsic rate of stem-cell division per year (irS), was also derived from the epiTOC2 algorithm (20). In all age groups, CRCs showed consistent and statistically significant AA by epiTOC (*p* < 2 × 10^−16^), epiTOC2 (*p <* 2 × 10^−16^) (**Figure 4C & D**), and also by irS (*p <* 2 × 10^−16^). The same pattern was also observed in the Hannum clock and PhenoAge, but not in the Horvath clock (**Supplementary Figure 4**).

To further exploit both mitotic clocks’ efficacy for estimating the cancer risk in non-cancerous normal tissues, we next applied both epiTOC and epiTOC2 to NM samples, with the primary aim to address whether AA or the intrinsic stem cell division rates differ between NMs from people with and without CRC, across the age groups. To eliminate the likely skew caused by the substantial AA in CRCs, we performed a linear regression by only including 129 and 231 NMs from people with and without CRC, respectively. This analysis was further adjusted for sex, as well as cell heterogeneities by incorporating the ratios of epithelial, fibroblast and immune cell compositions, which were estimated using the EpiDISH (42).

Both epiTOC and epiTOC2 predicted significant AA in NMs from EOCRC when compared with NMs from IOCRC and LOCRC (*p* = 0.003 for both epiTOC and epiTOC2; **Figure 5**). In NMs from healthy controls without CRCs, a reverse pattern was observed by both clocks (**Figure 5 A&B**), although this skew was likely induced by obtaining residuals from the linear regression, due to the skewed coefficients between two NM groups.

**Figure 5.**
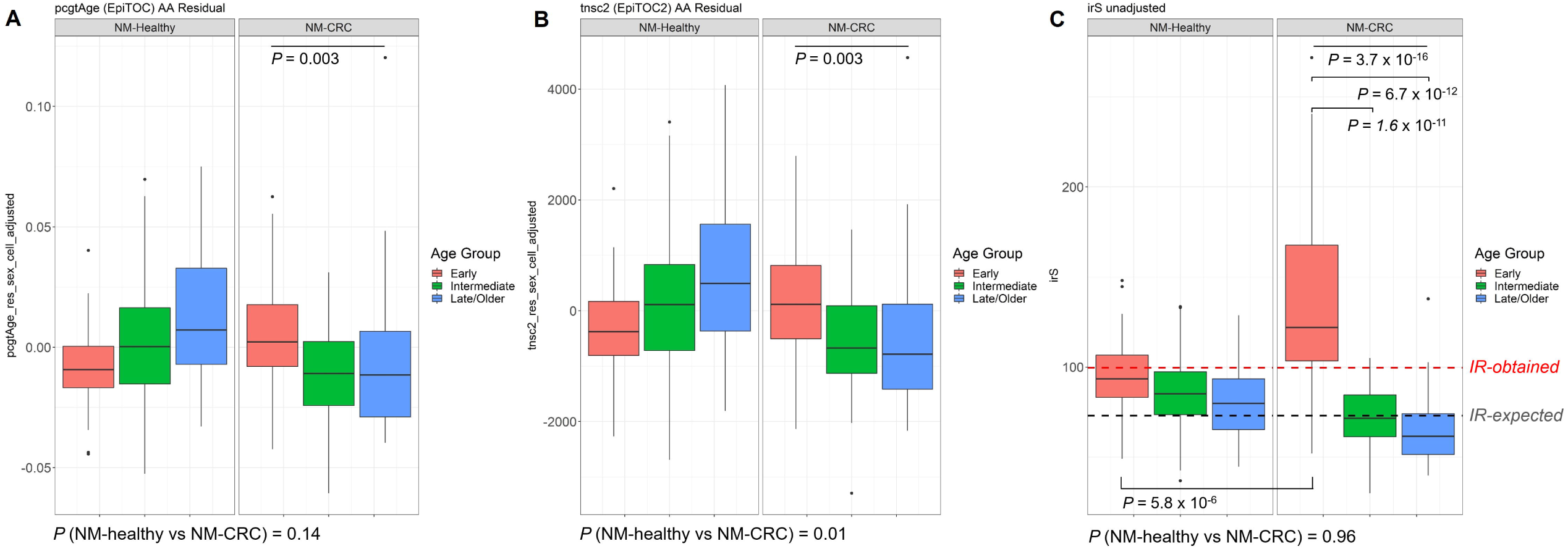
Boxplots showing the distributions of AA measured by epiTOC (**A**), epiTOC2 (**B**) and irS (**C**), in NMs from people without (“**NM-Healthy”**) and people with CRC (“**NM-CRC”**). AA was measured by calculating residuals from a linear regression analysis of DNAm age on chronological age for each individual. P-values between two tissue types (NM-Healthy vs. NM-CRC) were obtained by performing Wilcox tests whereas p-values between three age groups were obtained from Kruskal-Willis tests. In **(C)**, the red horizontal line denotes previously computed average *irS* (*IR*) in normal colonic mucusa (20) and the black horizontal line denotes an expected experimentally derived stem-cell division rate for colorectal tissue (21,43).

The irS was predicted to be significantly elevated in NM samples from the EOCRC group (*p =* 3.7 × 10^−16^, **Figure 5C**). The epiTOC2 clock measures the average lifetime intrinsic rate of stem-cell division per year for each sample, where this has been demonstrated to accurately distinguish precancerous normal colonic lesions (20). The irS, estimated by the epiTOC2 (and automatically adjusted for age), ranged from 37 (indicative of average 37 stem cell turnover per year) to 48 (mean = 86.7 ± 20 SD) for 231 NMs from healthy people, whereas for 129 NMs from people with CRCs ranged from 30 to 271 (mean = 96.4 ± 46 SD; *p*=0.96). A significantly higher irS was detected in NMs from EOCRC group when compared with IOCRC (*p* = 1.6 × 10^−11^) and LOCRC (*p* = 6.7 × 10^−12^) groups, indicating higher average stem-cell division rate in NMs from people with EOCRC (**Figure 5C**). Only the NM samples from the EOCRC group showed overall irS (mean = 135, median = 122) that were higher than previously observed by epiTOC2 (20) or biologically-determined stem-cell division rate (43) in normal colonic tissue. No significant differences in AA between the three age groups was observed in analyses of the three non-mitotic clocks (**Supplementary Figure 4**).

## Discussion

In this study, we examined the genome-wide DNAm changes associated with colorectal tumourigenesis across the full spectrum of age. We reported 88,358 consensus DMPs (20% of total CpGs tested) between the three CRC age groups, representing key somatic DNAm-related changes in tumourigenesis. Beyond these consensus DMPs, we identified 18,775 unique DMPs for EOCRC, which clustered into 234 DMRs, including the top-ranked DMRs related to the *TFAP2A* and *GSX1* genes. These genes have previously been reported to play a role in CRC tumourigenesis (44-46). Of note, *GSX1* methylation levels have been linked to obesity-associated CRC (44). Obesity is a recognised risk factor for EOCRC (47), which may support the hypothesis that risk factors such as obesity, may predispose to EOCRC by modulating DNAm of genes like *GSX1*. In addition, we found that the EOCRC DMRs were associated with a KEGG pathway of “Maturity onset diabetes of the young”. Our findings support the presence of unique DNAm molecular drivers and disrupted pathways in EOCRC.

Diabetes mellitus (DM) is a known risk factor for CRC (48). Although the rising trend of DM has been proposed to contribute to increasing EOCRC incidence (49), no particular association between DM and EOCRC has been shown, although only a limited number of reports are available (9,50). Maturity onset diabetes of the young (MODY) is a rare hereditary autoimmune condition that affects adolescents or young adults (51). Like other forms of diabetes, MODY is linked to obesity and hyperglycaemia (51), which are known causative factors for intestinal inflammation and also linked to the gut microbiome dysbiosis (52). Whilst no information about this atypical form of diabetes was collected in our cohorts, MODY-related genes have been reported to be epigenetically dysregulated in CRCs (of no particular onset/subtype) (53), and hypermethylated with ageing in people at high risk for metabolic syndromes (54), warranting further investigation as a specific risk factor for EOCRC.

The adverse role of ageing-associated methylomic drift in predisposing people to CRC has become clear and is now postulated to be the primary mechanism for explaining ageing-related CRC risk (13,14,55). Recent data utilising the high-density methylation arrays have specifically predicted a long sojourn time from early mostly benign ageing-associated methylation changes to the formation of premalignant cells and suggested that early methylation changes can be traced back several decades (14). We hypothesised that EOCRC shows an accelerated ageing process with a more rapid transition time from benign to malignant cells and that this would be reflected in an increase in cancer-associated drift CpGs in EOCRC and/or by measures of AA in the NM of people with EOCRC. In NMs, without longitudinal sampling, our data was unable to directly measure methylomic drift rates within each individual with EOCRC. However, we showed that a high proportion (>58%) of CpGs associated with methylomic drift are aberrantly methylated in CRC tumours of all ages. Remarkably, DNAm aberrations at these cancer-associated drift-CpGs were also widely affected in EOCRC and this was as extensive as in IOCRC and LOCRC, which suggests that DNAm of cancer-associated drift CpGs is essential for CRC tumourigenesis at all ages, even in young people at risk. Our data show that EOCRCs rapidly acquire extensive DNAm aberration at these drift-CpGs without needing a life-long accumulation of cancer-related methylomic drift at these drift CpGs.

Cancer (and especially cancer of non-hereditary origin) is a phenomenon related to cumulative DNAm aberrations including those of ageing-associated methylomic drift (13). Methylomic drift, defined as a “gradual change away from baseline (i.e. birth or conception)”, is postulated to arise due to the imperfect epigenetic maintenance machinery (13). Though the DNAm changes associated with methylomic drift are estimated to be subtle over shorter period of time, tissues that are highly proliferative and directly exposed to exogeneous factors (e.g. diet, microbiota) such as colonic epithelium may have a slightly faster drift than other tissues (13,56). We hypothesise that, in our EOCRC tumours, a defective epigenetic machinery or adverse environmental exposures may have synergistically contributed to this rapid transformation in relatively shorter periods of time. Further studies identifying causative factors and predictive biomarkers for early detection may help prevent sporadic EOCRCs. A longitudinal sampling of NMs from the same individuals at higher risk of CRC could be used to estimate “endogenous” methylomic drift rate for each individual and tease out the effect of specific genetic factors and environmental exposures to this cancer-associated drift.

In NMs from people with EOCRC, we demonstrated significant AA, when compared with NMs from people with IOCRC or LOCRC, or NMs from young healthy people without CRC, as measured using mitotic-based DNAm clocks. By utilising two recently developed mitotic-based DNAm clocks, we showed that NMs from EOCRC group exhibit the total mitotic stem cell division numbers that were comparable to NMs of IOCRC and LOCRC groups, suggesting an elevated mitotic rate per given years of age in our EOCRC group. The irS tool estimates the average intrinsic rate of stem cell division per year. We found further support that accelerated ageing in NMs from people with EOCRC, evidenced as being the only group showing the mean irS exceeding the expected stem-cell division rates in healthy colonic mucosa (37,55) and significantly different from NMs of IOCRC, LOCRC or NMs of people without CRC.

The increased mitotic rate and DNAm errors that arise from this has been proposed to be a major determinant pf cancer risk (13,20,21,57,58) and here, we have shown accelerated ageing in non-cancerous NMs from people who developed EOCRC, which may constitute an important risk predictor for EOCRC development. The epiTOC and epiTOC2 mitotic-based clocks used in this study have been proven to accurately predict an increase in cell proliferation, a phenomenon associated with inflammatory and precancerous tissues, various cancer types and even in the buccal epithelial cells of smokers (20,21), demonstrating its potential as a cancer-risk estimator in pre-diagnostic tissue samples. It is likely that the mitotic rate of individual tissue results from an interplay between intrinsic and extrinsic factors such as inflammation, environmental and the gut microbiome (13,58), and we hypothesise that disruption in these factors may be reflected in the accelerated (mitotic) age in NMs from EOCRC. Whilst this hypothesis remains plausible, our study was not designed to directly assess such association. Future studies investigating this potential interrelation between specific in/extrinsic factors and disruptions in the mitotic rate could have the particular importance for EOCRC prevention.

CpG island methylator phenotype (CIMP)-high or CIMP-positive CRC represents ∼20% of CRC and is associated with the serrated pathway of tumourigenesis and an older age at diagnosis (59). This subtype of CRC is characterised by widespread and concordant DNA methylation changes (59). In our study, CIMP-positive CRCs represented only a small proportion of all CRCs (we excluded CRCs identified by *MLH1* promoter methylation and loss of MLH1/PMS2 protein expression (MMR-deficient)). In addition, the CA-drift CpGs showed minimal overlap with commonly assessed CIMP-related CpGs, therefore, CIMP is unlikely to be the underlying driver of the CA-drift CpGs reported in this study.

This study has some limitations. We utilised a public dataset of “healthy” NM therefore, some inherent technical variations (e.g. batch effect, tissue source) is not unexpected. A possible inconsistency due to tissue storage or batch effects cannot be ruled out. Ideally, a set of colonic NM samples collected longitudinally from the same individuals would serve as a perfect archetype to investigate this though we are not aware of such cohort. Another limitation is the lack of gene expression data to assess the functional implications of the methylation changes.

## Conclusion

Our study represents one of the largest genome-wide DNAm studies on EOCRC. Utilising participants recruited via two internationally unique cohorts, we have generated methylome data for 769 non-hereditary MMR-proficient CRCs including 97 tumour samples (with 41 matched tumour-normal pairs) from people with EOCRC. Several novel findings from this study include: (i) extensive DNAm alterations in all CRCs including EOCRCs, indicative of the essential role DNAm plays in CRC development at all ages, (ii) the identification of unique DNAm-related changes in EOCRC including at the *TFAP2A* and *GSX1* genes and genes associated with MODY, (iii) EOCRC tumours have extensive DNAm aberrations at ageing-associated CpGs or CA-drift CpGs without requiring a decades-long accumulation of methylation aberrations, which has been postulated to be essential in conventional CRC development that commonly occurs at an older age (13,14), and (iv) AA related to accelerated mitotic stem-cell division rates (i.e. accelerated tissue ageing) in NMs from people with EOCRC. These findings suggest that tumourigenesis in EOCRC is molecularly unique. Further studies investigating the cause of these DNAm changes will help elucidate the determinants of the increasing incidence of EOCRC and aid in the identification of predictive biomarkers that can lead to new prevention strategies in young people who are at higher risk of CRC.

## Data Availability

The datasets of the current study are available from the corresponding author on reasonable request.

## ABBREVIATIONS

AA: Age acceleration
ACCFR: Australian Colon Cancer Family Registry
AgeDx: Age of diagnosis
CIMP: CpG Island Methylator Phenotype
CpG: Cytosine-(phosphate)-Guanine dinucleotide
CRC: Colorectal cancer
DNAm: DNA methylation
DMP: Differentially methylated probe
DMR: Differentially methylated region
Dx: diagnosis
EOCRC: Early-onset colorectal cancer
IOCRC: Intermediate-onset colorectal cancer
LOCRC: Late-onset colorectal cancer
FDR: False discovery rate
MMR: Mismatch repair
MSI: Microsatellite instable
MSS: Microsatellite stable
MCCS: Melbourne Collaborative Cohort Study
NM: Normal colonic mucosa
KEGG: Kyoto Encyclopedia of Genes and Genomes

## Funding/Support

Melbourne Collaborative Cohort Study (MCCS) cohort recruitment was funded by VicHealth and Cancer Council Victoria. The MCCS was further augmented by Australian National Health and Medical Research Council grants 209057, 396414 and 1074383 and by infrastructure provided by Cancer Council Victoria. Cases and their vital status were ascertained through the Victorian Cancer Registry and the Australian Institute of Health and Welfare, including the National Death Index and the Australian Cancer Database (ACD). New South Wales (NSW) cancer registry data were obtained via the ACD with the assistance of the NSW Ministry of Health.

The Colon Cancer Family Registry (CCFR)was supported by the National Cancer Institute of the National Institutes of Health under Award Number U01CA167551 and through a cooperative agreement with the Australasian Colorectal Cancer Family Registry (NCI/NIH U01 CA074778 and U01/U24 CA097735) and by the Victorian Cancer Registry, Australia. This research was performed under CCFR approved project C-AU-0312-01.

DDB is supported by a NHMRC R.D. Wright Career Development Fellowship, a NHMRC Emerging Leadership Investigator grant and by funding from the University of Melbourne Research at Melbourne Accelerator Program (R@MAP). PG is supported by an Australian Government Research Training Program Scholarship. AKW is a NHMRC Career Development Fellow. JLH is a NHMRC Senior Principal Research Fellow. MAJ is a NHMRC Senior Research Fellow.

## Acknowledgements

We thank the participants and staff from the Colon Cancer Family Registry (CCFR) and the Melbourne Collaboratory Cohort (MCCS). We especially thank Maggie Angelakos, Samantha Fox, Allyson Templeton for facilitating CCFR resources and supporting this study. We also thank members of the Colorectal Oncogenomics Group (The University of Melbourne) and the Precision Medicine (Monash University) for supporting this study.

## Contributors

JEJ and DDB conceived the original idea and designed the study. CR, IMW, DE, FAM, AKW, JLH, MAJ, RLM, GGG, MCS and DDB contributed to the acquisition of data. AKW, JLH, MAJ and DDB are investigators for the ACCFR. DE, RLM, GGG and MCS are investigators for the MCCS. JEJ, MC, EMW and SGP performed the laboratory experiment and contributed to generating DNA methylation data. The bioinformatics analyses were performed by JEJ in support of KH, PG and PAD. The statistical analyses were performed and reviewed by JEJ, AKW, PAD, HJ, JHP, MAJ, RLM and DDB. JEJ and DDB prepared figures/tables and the first draft of the manuscript. All authors read and provided critical revisions to the manuscript and approved the final manuscript.

## Competing interests

The authors declare no competing interests.

---

**Supplementary Figure 1.**
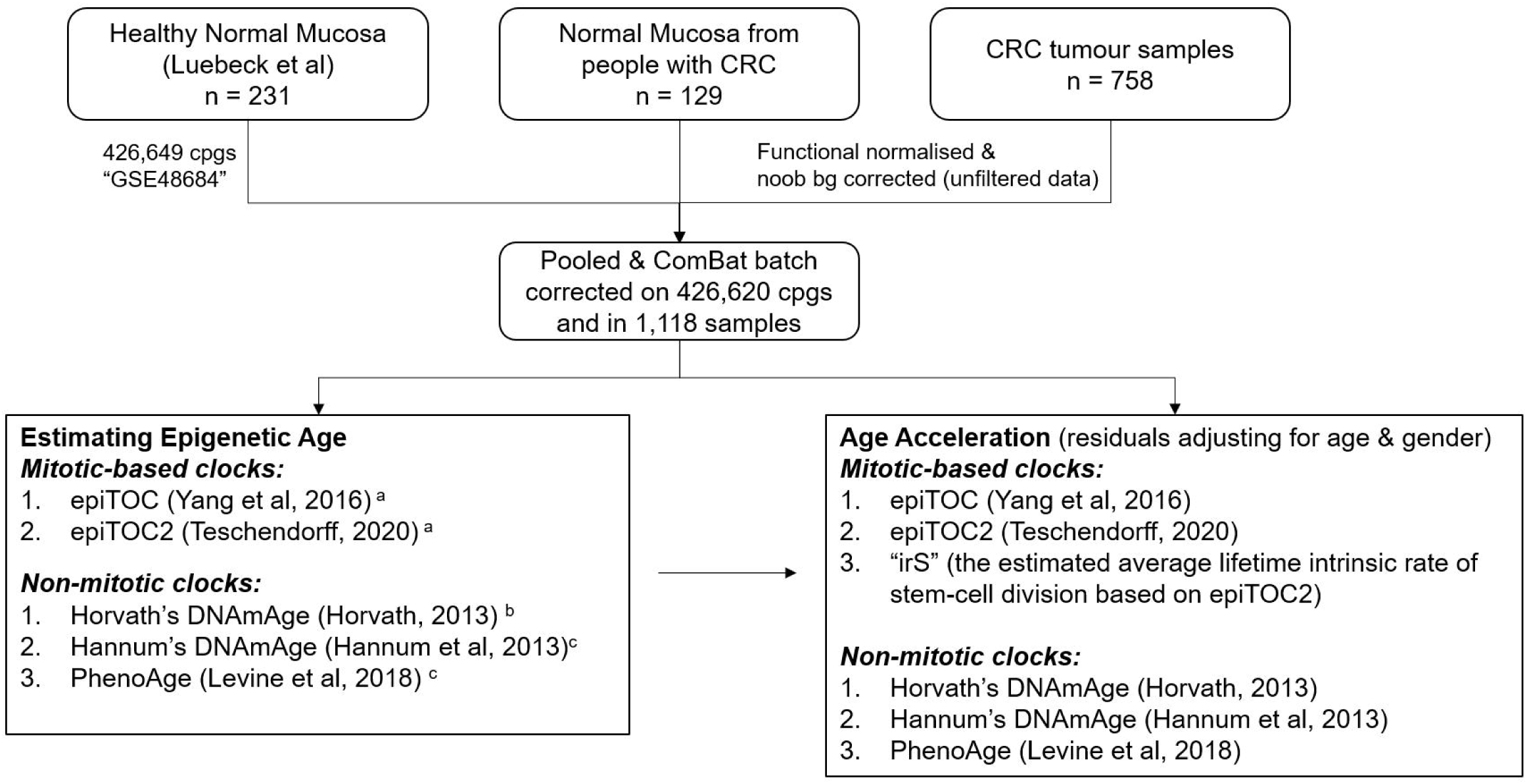
A flow diagram showing the data processing steps for deriving DNAm-age from 231 NMs (without CRC), 129 NMs from people with CRC and 758 CRC tumour samples. ^a^ as described in Teschendorff 2020, ^b^ using the online calculator (http://dnamage.genetics.ucla.edu/new), and ^c^ using the *ENmix* Bioconductor package.

**Supplementary Figure 2.**
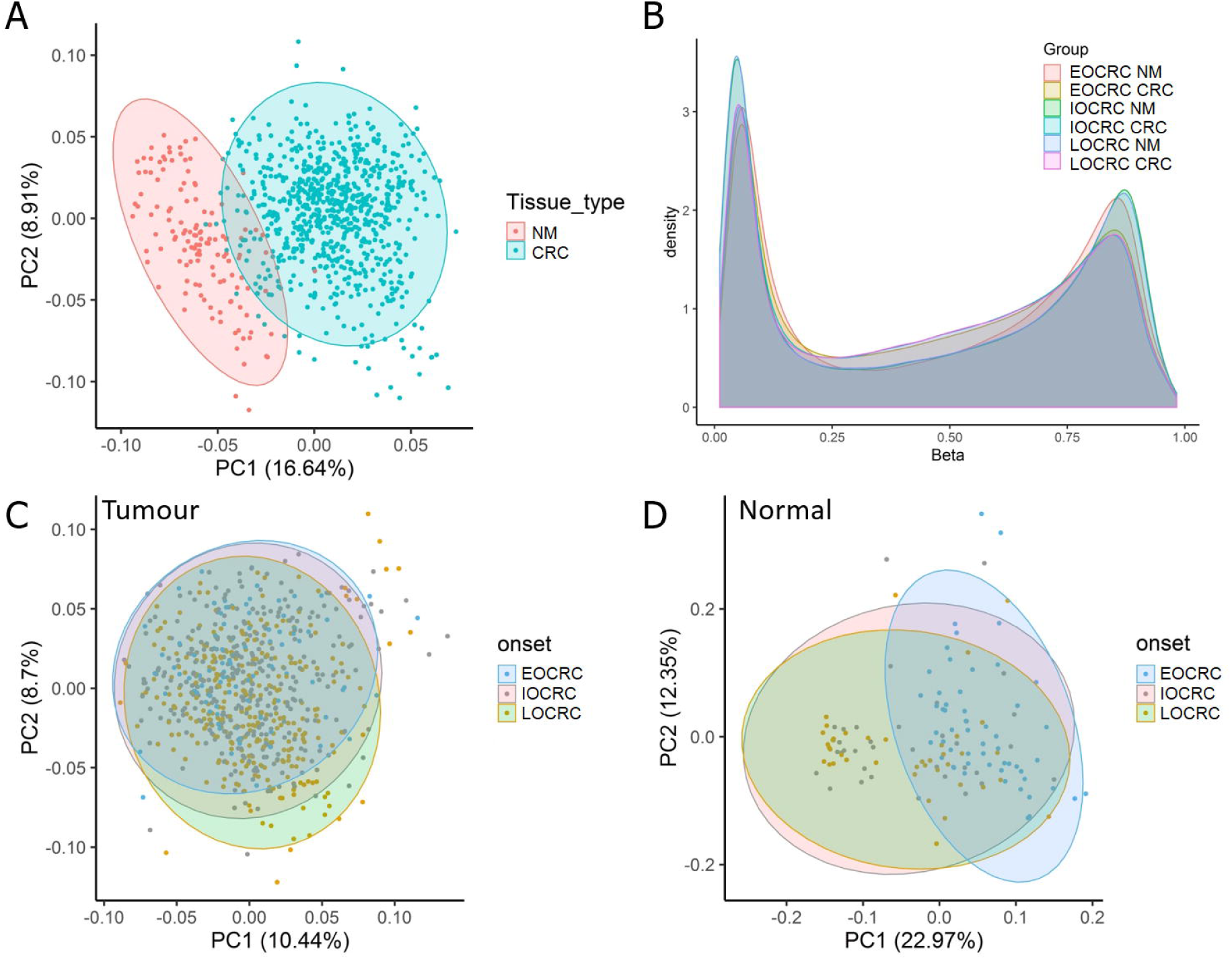
PCA plot (**A**) and β methylation density plot (**B**) showing overall DNA methylation similarities between samples. Individual samples are coloured by tissue type (tumour/normal mucosa). PCA plots for tumour (**C**) and normal colonic mucosa samples only (**D**).

**Supplementary Figure 3.**
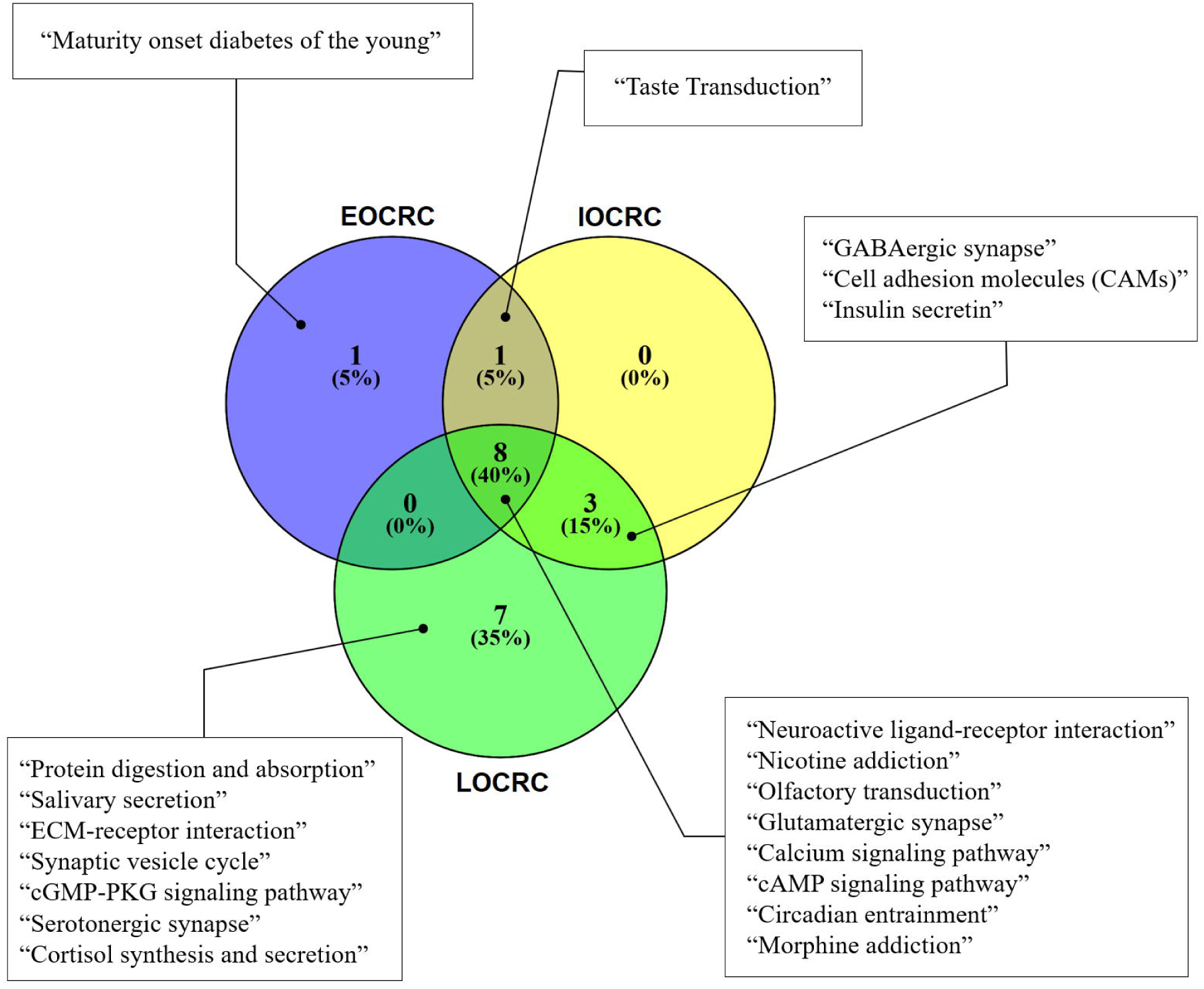
KEGG pathways associated with differentially methylated genes of EOCRC, IOCRC and LOCRC. Numbers of KEGG pathways unique to each CRC onset groups as well as numbers of overlapping pathways are shown.

**Supplementary Figure 4.**
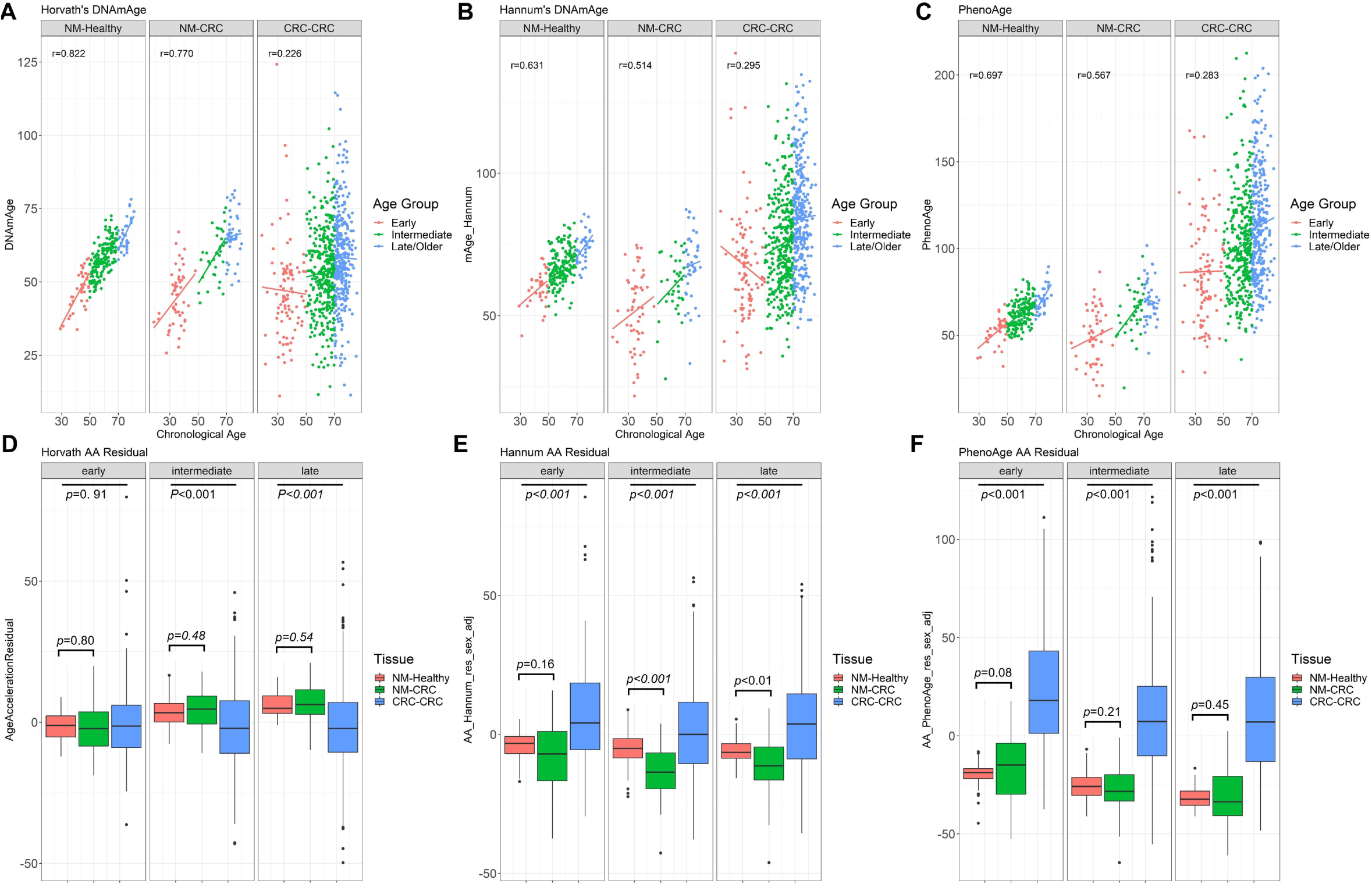
DNA methylation age and AA measured using non-mitotic epigenetic clocks. Scatterplots showing correlations between DNAm-derived ages using the Horvath (**A**) and Hannum (**B**) clocks, and PhenoAge (**C**) for 231 NMs from healthy people (“NM-Healthy”), 129 NMs from people with CRC (“NM-CRC”) and 758 CRC tumour samples (“CRC-CRC”). Age Acceleration (AA) were estimated by obtaining residuals from a linear regression of DNAm-ages on chronological age, adjusting for sex. Boxplots showing the AA distributions, as estimated by the Horvath (**D**) and Hannum (**E**) clocks, and PhenoAge (**F**). *P-*values were computed by Wilcoxon Rank Sum test for two groups (NM-Healthy vs NM-CRC) or by Kruskal-Wallis Rank Sum test for three groups (NM-Healthy vs NM-CRC vs CRC-CRC).

## Notes

### Competing Interest Statement

The authors have declared no competing interest.

## References

1. Brenner H, Chen C. The colorectal cancer epidemic: challenges and opportunities for primary, secondary and tertiary prevention. British journal of cancer 2018;119:785–92

2. Brenner H, Kloor M, Pox CP. Colorectal cancer. Lancet 2014;383:1490–502

3. Ahnen DJ, Wade SW, Jones WF, Sifri R, Mendoza Silveiras J, Greenamyer J, et al. The increasing incidence of young-onset colorectal cancer: a call to action. Mayo Clin Proc 2014;89:216–24

4. Pearlman R, Frankel WL, Swanson B, Zhao W, Yilmaz A, Miller K, et al. Prevalence and Spectrum of Germline Cancer Susceptibility Gene Mutations Among Patients With Early-Onset Colorectal Cancer. JAMA oncology 2017;3:464–71

5. Stoffel EM, Koeppe E, Everett J, Ulintz P, Kiel M, Osborne J, et al. Germline Genetic Features of Young Individuals With Colorectal Cancer. Gastroenterology 2017

6. Baylin SB, Jones PA. Epigenetic Determinants of Cancer. Cold Spring Harbor perspectives in biology 2016;8

7. Walters RJ, Williamson EJ, English DR, Young JP, Rosty C, Clendenning M, et al. Association between hypermethylation of DNA repetitive elements in white blood cell DNA and early-onset colorectal cancer. Epigenetics : official journal of the DNA Methylation Society 2013;8:748–55

8. Antelo M, Balaguer F, Shia J, Shen Y, Hur K, Moreira L, et al. A high degree of LINE-1 hypomethylation is a unique feature of early-onset colorectal cancer. PloS one 2012;7:e45357

9. Rosato V, Bosetti C, Levi F, Polesel J, Zucchetto A, Negri E, et al. Risk factors for young-onset colorectal cancer. Cancer causes & control : CCC 2013;24:335–41

10. Belshaw NJ, Pal N, Tapp HS, Dainty JR, Lewis MP, Williams MR, et al. Patterns of DNA methylation in individual colonic crypts reveal aging and cancer-related field defects in the morphologically normal mucosa. Carcinogenesis 2010;31:1158–63

11. Luo Y, Yu M, Grady WM. Field cancerization in the colon: a role for aberrant DNA methylation? Gastroenterology report 2014;2:16–20

12. Shen L, Kondo Y, Rosner GL, Xiao L, Hernandez NS, Vilaythong J, et al. MGMT promoter methylation and field defect in sporadic colorectal cancer. Journal of the National Cancer Institute 2005;97:1330–8

13. Issa JP. Aging and epigenetic drift: a vicious cycle. The Journal of clinical investigation 2014;124:24–9

14. Luebeck GE, Hazelton WD, Curtius K, Maden SK, Yu M, Carter KT, et al. Implications of epigenetic drift in colorectal neoplasia. Cancer research 2018

15. Noreen F, Roosli M, Gaj P, Pietrzak J, Weis S, Urfer P, et al. Modulation of age-and cancer-associated DNA methylation change in the healthy colon by aspirin and lifestyle. Journal of the National Cancer Institute 2014;106

16. Hannum G, Guinney J, Zhao L, Zhang L, Hughes G, Sadda S, et al. Genome-wide methylation profiles reveal quantitative views of human aging rates. Molecular cell 2013;49:359–67

17. Horvath S. DNA methylation age of human tissues and cell types. Genome biology 2013;14:R115

18. Horvath S, Raj K. DNA methylation-based biomarkers and the epigenetic clock theory of ageing. Nature reviews Genetics 2018;19:371–84

19. Levine ME, Lu AT, Quach A, Chen BH, Assimes TL, Bandinelli S, et al. An epigenetic biomarker of aging for lifespan and healthspan. Aging 2018;10:573–91

20. Teschendorff AE. A comparison of epigenetic mitotic-like clocks for cancer risk prediction. Genome Med 2020;12:56

21. Yang Z, Wong A, Kuh D, Paul DS, Rakyan VK, Leslie RD, et al. Correlation of an epigenetic mitotic clock with cancer risk. Genome biology 2016;17:205

22. Zhou W, Dinh HQ, Ramjan Z, Weisenberger DJ, Nicolet CM, Shen H, et al. DNA methylation loss in late-replicating domains is linked to mitotic cell division. Nature genetics 2018;50:591–602

23. Jenkins MA, Win AK, Templeton AS, Angelakos MS, Buchanan DD, Cotterchio M, et al. Cohort Profile: The Colon Cancer Family Registry Cohort (CCFRC). International journal of epidemiology 2018

24. Newcomb PA, Baron J, Cotterchio M, Gallinger S, Grove J, Haile R, et al. Colon Cancer Family Registry: an international resource for studies of the genetic epidemiology of colon cancer. Cancer epidemiology, biomarkers & prevention : a publication of the American Association for Cancer Research, cosponsored by the American Society of Preventive Oncology 2007;16:2331–43

25. Milne RL, Fletcher AS, MacInnis RJ, Hodge AM, Hopkins AH, Bassett JK, et al. Cohort Profile: The Melbourne Collaborative Cohort Study (Health 2020). International journal of epidemiology 2017;46:1757–i

26. Buchanan DD, Clendenning M, Rosty C, Eriksen SV, Walsh MD, Walters RJ, et al. Tumor testing to identify lynch syndrome in two Australian colorectal cancer cohorts. Journal of gastroenterology and hepatology 2017;32:427–38

27. Wong EM, Joo JE, McLean CA, Baglietto L, English DR, Severi G, et al. Tools for translational epigenetic studies involving formalin-fixed paraffin-embedded human tissue: applying the Infinium HumanMethyation450 Beadchip assay to large population-based studies. BMC research notes 2015;8:543

28. Aryee MJ, Jaffe AE, Corrada-Bravo H, Ladd-Acosta C, Feinberg AP, Hansen KD, et al. Minfi: a flexible and comprehensive Bioconductor package for the analysis of Infinium DNA methylation microarrays. Bioinformatics 2014;30:1363–9

29. Fortin JP, Labbe A, Lemire M, Zanke BW, Hudson TJ, Fertig EJ, et al. Functional normalization of 450k methylation array data improves replication in large cancer studies. Genome biology 2014;15:503

30. Triche TJ, Jr., Weisenberger DJ, Van Den Berg D, Laird PW, Siegmund KD. Low-level processing of Illumina Infinium DNA Methylation BeadArrays. Nucleic acids research 2013;41:e90

31. Du P, Zhang X, Huang CC, Jafari N, Kibbe WA, Hou L, et al. Comparison of Beta-value and M-value methods for quantifying methylation levels by microarray analysis. BMC bioinformatics 2010;11:587

32. Ritchie ME, Phipson B, Wu D, Hu Y, Law CW, Shi W, et al. limma powers differential expression analyses for RNA-sequencing and microarray studies. Nucleic acids research 2015;43:e47

33. Peters TJ, Buckley MJ, Statham AL, Pidsley R, Samaras K R VL, et al. De novo identification of differentially methylated regions in the human genome. Epigenetics & chromatin 2015;8:6

34. Wickham H. ggplot2: Elegant Graphics for Data Analysis. Springer-Verlag New York; 2016.

35. Phipson B, Maksimovic J, Oshlack A. missMethyl: an R package for analyzing data from Illumina’s HumanMethylation450 platform. Bioinformatics 2016;32:286–8

36. Adams SV, Newcomb PA, Burnett-Hartman AN, Wurscher MA, Mandelson M, Upton MP, et al. Rare circulating microRNAs as biomarkers of colorectal neoplasia. PloS one 2014;9:e108668

37. Barault L, Amatu A, Siravegna G, Ponzetti A, Moran S, Cassingena A, et al. Discovery of methylated circulating DNA biomarkers for comprehensive non-invasive monitoring of treatment response in metastatic colorectal cancer. Gut 2018;67:1995–2005

38. Leek JT, Johnson WE, Parker HS, Jaffe AE, Storey JD. The sva package for removing batch effects and other unwanted variation in high-throughput experiments. Bioinformatics 2012;28:882–3

39. Xu Z, Niu L, Li L, Taylor JA. ENmix: a novel background correction method for Illumina HumanMethylation450 BeadChip. Nucleic acids research 2016;44:e20

40. McEwen LM, Jones MJ, Lin DTS, Edgar RD, Husquin LT, MacIsaac JL, et al. Systematic evaluation of DNA methylation age estimation with common preprocessing methods and the Infinium MethylationEPIC BeadChip array. Clinical epigenetics 2018;10:123

41. Wang T, Maden SK, Luebeck GE, Li CI, Newcomb PA, Ulrich CM, et al. Dysfunctional epigenetic aging of the normal colon and colorectal cancer risk. Clinical epigenetics 2020;12:5

42. Zheng SC, Breeze CE, Beck S, Teschendorff AE. Identification of differentially methylated cell types in epigenome-wide association studies. Nat Methods 2018;15:1059–66

43. Tomasetti C, Vogelstein B. Cancer etiology. Variation in cancer risk among tissues can be explained by the number of stem cell divisions. Science 2015;347:78–81

44. Crujeiras AB, Morcillo S, Diaz-Lagares A, Sandoval J, Castellano-Castillo D, Torres E, et al. Identification of an episignature of human colorectal cancer associated with obesity by genome-wide DNA methylation analysis. International journal of obesity 2019;43:176–88

45. Graham RP, Shrestha B, Caron BL, Smyrk TC, Grogg KL, Lloyd RV, et al. Islet-1 is a sensitive but not entirely specific marker for pancreatic neuroendocrine neoplasms and their metastases. The American journal of surgical pathology 2013;37:399–405

46. Ropponen KM, Kellokoski JK, Pirinen RT, Moisio KI, Eskelinen MJ, Alhava EM, et al. Expression of transcription factor AP-2 in colorectal adenomas and adenocarcinomas; comparison of immunohistochemistry and in situ hybridisation. J Clin Pathol 2001;54:533–8

47. Araghi M, Fidler MM, Arnold M, Jemal A, Bray F, Soerjomataram I. The Future Burden of Colorectal Cancer Among US Blacks and Whites. Journal of the National Cancer Institute 2018;110:791–3

48. Yuhara H, Steinmaus C, Cohen SE, Corley DA, Tei Y, Buffler PA. Is diabetes mellitus an independent risk factor for colon cancer and rectal cancer? Am J Gastroenterol 2011;106:1911–21; quiz 22

49. Young JP, Win AK, Rosty C, Flight I, Roder D, Young GP, et al. Rising incidence of early-onset colorectal cancer in Australia over two decades: report and review. Journal of gastroenterology and hepatology 2015;30:6–13

50. Gausman V, Dornblaser D, Anand S, Hayes RB, O’Connell K, Du M, et al. Risk Factors Associated With Early-Onset Colorectal Cancer. Clin Gastroenterol Hepatol 2019

51. Anik A, Catli G, Abaci A, Bober E. Maturity-onset diabetes of the young (MODY): an update. J Pediatr Endocrinol Metab 2015;28:251–63

52. Thaiss CA, Levy M, Grosheva I, Zheng D, Soffer E, Blacher E, et al. Hyperglycemia drives intestinal barrier dysfunction and risk for enteric infection. Science 2018;359:1376–83

53. Liu J, Li H, Sun L, Wang Z, Xing C, Yuan Y. Aberrantly methylated-differentially expressed genes and pathways in colorectal cancer. Cancer Cell Int 2017;17:75

54. Ali O, Cerjak D, Kent JW, Jr., James R, Blangero J, Carless MA, et al. An epigenetic map of age-associated autosomal loci in northern European families at high risk for the metabolic syndrome. Clinical epigenetics 2015;7:12

55. Ahuja N, Li Q, Mohan AL, Baylin SB, Issa JP. Aging and DNA methylation in colorectal mucosa and cancer. Cancer research 1998;58:5489–94

56. Maegawa S, Hinkal G, Kim HS, Shen L, Zhang L, Zhang J, et al. Widespread and tissue specific age-related DNA methylation changes in mice. Genome research 2010;20:332–40

57. Nejman D, Straussman R, Steinfeld I, Ruvolo M, Roberts D, Yakhini Z, et al. Molecular rules governing de novo methylation in cancer. Cancer research 2014;74:1475–83

58. Yu M, Hazelton WD, Luebeck GE, Grady WM. Epigenetic Aging: More Than Just a Clock When It Comes to Cancer. Cancer research 2020;80:367–74

59. Weisenberger DJ, Liang G, Lenz HJ. DNA methylation aberrancies delineate clinically distinct subsets of colorectal cancer and provide novel targets for epigenetic therapies. Oncogene 2018;37:566–77

